# The application of Large Language Models to the phenotype-based prioritization of causative genes in rare disease patients

**DOI:** 10.1101/2023.11.16.23298615

**Authors:** Şenay Kafkas, Marwa Abdelhakim, Azza Althagafi, Sumyyah Toonsi, Malak Alghamdi, Paul N. Schofield, Robert Hoehndorf

## Abstract

Computational methods for identifying gene–disease associations can use both genomic and phenotypic information to prioritize genes and variants that may be associated with genetic diseases. Phenotype-based methods commonly rely on comparing phenotypes observed in a patient with a database of genotype-to-phenotype associations using a measure of semantic similarity, and are primarily limited by the quality and completeness of this database as well as the quality of phenotypes assigned to a patient. Genotype-to-phenotype associations used by these methods are largely derived from literature and coded using phenotype ontologies. Large Language Models (LLMs) have been trained on large amounts of text and have shown their potential to answer complex questions across multiple domains. Here, we demonstrate that LLMs can prioritize disease-associated genes as well, or better than, dedicated bioinformatics methods relying on calculated phenotype similarity. The LLMs use only natural language information as background knowledge and do not require ontology-based phenotyping or structured genotype-to-phenotype knowledge. We use a cohort of undiagnosed patients with rare diseases and show that LLMs can be used to provide diagnostic support that helps in identifying plausible candidate genes.

## 1 Introduction

Rare diseases individually affect a small number of people in the population; yet, despite their low prevalence, the collective impact of rare diseases on global public health is substantial, affecting millions of individuals worldwide [1]. However, diagnosing rare diseases is particularly challenging due to the often small number of affected individuals with a specific disorder and the variation in disease presentation between patients. Consequently, patients with rare diseases often endure a diagnostic odyssey, undergoing numerous tests and consultations over years before receiving an accurate diagnosis.

Most rare diseases have a Mendelian basis and are the result of variation in one or at most a small number of genes [2]. Next-generation sequencing (NGS) has revolutionized the diagnosis of Mendelian diseases and whole exome and genome sequencing can identify genetic variants in individuals which may cause the disorder; the number of these variants ranges from around 20-40,000 in the whole exome to over a million in whole genome sequencing [3, 4].

The challenge lies in identifying the genetic variant or variants that lead to the disease in a particular individual. Although sequencing has made a huge impact on disease identification, only about 50% of patients end up with molecular diagnosis [5]. A typical genome will contain 100-200 genes with protein-truncating variants and around 10,000 to 12,000 sites with variants causing changes in peptide sequence. Estimates suggest that an individual carries around 100 loss-of-function (LOF) alleles with around 20 completely inactivated [6]. In a recent large-scale study of European populations, most individuals were assessed to have 2-5 autosomal recessive pathogenic or likely pathogenic variants in known Mendelian disease genes, most of which had an allele frequency of less than 0.001 [7]. Most individuals within a population, therefore, carry at least one disease allele for any recessive disease, and many potentially pathogenic variants in genes where the variant either does not meet the loss-of-function threshold or where the gene is not already disease-associated. In a study of a large number of whole exome sequences, 3,230 genes highly intolerant to a loss-of-function were identified, 72% of which had no established human disease phenotype [8]. Prioritising which of the potentially pathogenic variants cause disease in a particular patient is therefore a complex task, especially given phenotypic variation and the action of modifiers.

Several methods are commonly employed to reduce the number of variants to consider, including the mode of inheritance, the frequency of observing the variant within different populations, and the functional impact a variant has on the function of a gene product. However, even after these filters have been applied, there are often still several variants left that need to be considered and evaluated [9–11].

In response to this challenge, various methods have been developed to predict whether a variant is pathogenic or will alter the function of a gene product. These methods include rule-based methods and machine learning methods [12, 13]. However, none of these methods is entirely accurate, and, moreover, multiple genes may suffer from a total loss of function without any abnormal phenotypic effects [14, 15]. The further challenge is therefore to identify which of the candidate variants is responsible for the phenotype observed.

Phenotype-based methods rank genes or genotypes based on whether they are likely to cause the phenotypes observed in a patient. Phenotype-based methods require that phenotypes are specified in a formal language, usually based on the Human Phenotype Ontology (HPO) [16]. The HPO is an ontology that provides over 17,000 standardized phenotype descriptions and several resources have been developed around the HPO, including large genotype-to-phenotype databases.

Phenotype-based methods for prioritizing genes or genotypes typically compare the phenotypes observed in a patient with the phenotypes in a genotype-to-phenotype database [17]. Because phenotypes may be variable, measures of semantic similarity are usually employed; an ontology-based semantic similarity measure uses the knowledge in an ontology to define a similarity measure between entities associated with ontology terms [18, 19]. Phenotype-based methods are therefore crucially dependent on three different resources: a phenotype ontology that provides domain knowledge by structuring and relating phenotype terms; a genotype-to-phenotype database; and a semantic similarity measure. Phenotype ontologies are manually curated and rely on domain expertise as well as knowledge of formal semantics [20]; genotype-to-phenotype databases are created from literature or large-scale experiments and may be incomplete or noisy [16]; and, although a large number of semantic similarity measures have been developed, they have different biases which makes them a challenge to apply consistently [21].

Large Language Models (LLMs) are now available that are trained in a self-supervised manner on large text corpora [22]. LLMs trained on large text corpora can also perform a large variety of different tasks and can further be fine-tuned to follow instructions or “prompts” [23]. They have been applied successfully to a wide range of tasks, including clinical question answering [24], medical reasoning, record keeping, and patient facing interactions [25]. We explore the potential of LLMs to overcome the limitations of formal phenotype similarity-based methods for ranking genes or variants. Their training on large text corpora may allow them to access the same, or more, information as is used in constructing genotype-to-phenotype databases and the phenotype ontologies, and potentially to capture semantic relationships between concepts [26]. Consequently, they may also be able to estimate semantic similarity as well as, or better than, ontology-dependent similarity measures.

Here, we apply and evaluate three LLMs, GPT-3.5-turbo [27], GPT-4 [28], and Falcon180B [29], in ranking genes based on clinically observed phenotypes, and we include the LLMs as part of a workflow that identifies disease-causing variants in whole exome or whole genome sequencing data. For our evaluation, we use three different synthetic datasets as well as one dataset of patients with undiagnosed genetic diseases, and we demonstrate by direct comparison to state of the art methods that LLMs can improve phenotype-based ranking of genes over all state of the art methods. Furthermore, interactions with LLMs can be used to generate explanations for ranking genes and can also be used to refine ranking results, demonstrating that LLMs have the potential to be used as diagnostic assistants. However, we also observed several cases of “hallucinations” and other biases, which need to be addressed before LLMs can be used reliably in a clinical context.

## 2 Materials and Methods

### 2.1 Datasets Used

We used three benchmark datasets to conduct our experiments. The first dataset, GPCards [30], is a manually curated dataset of genotype–phenotype associations. We randomly selected 50 variants from distinct genes along with their corresponding clinical phenotypes from GPCards. The phenotypes in GPCards are represented as natural language terms and do not rely on a structured vocabulary or ontology. We use the GPCards dataset to develop prompts and assess the performance of the LLMs on different prompts.

The second dataset is the October 2023 release of ClinVar [31] a publicly accessible database detailing genomic variations and their connections to disease. We focused particularly on the new variants included in ClinVar between July 2, 2023, and October 7, 2023. From this subset of data, we randomly selected 100 variants, each from a different gene, associated with diseases in Online Mendelian Inheritance in Man (OMIM) [32] We identified the phenotypes corresponding to the OMIM disease using the HPO database [33] accessed on October 8, 2023.

The third dataset is the Phenotype-Associated Variants in Saudi Arabia (PAVS) database [34], a public database of genotype–phenotype relations identified in Saudi individuals. PAVS combines a collection of clinically validated pathogenic variants with manually curated variants specific to the Saudi population, each accompanied by its associated phenotypes mapped to HPO codes. We used the PAVS dataset to compare LLMs with ontology-based gene prioritization methods. The phenotypes in PAVS also correspond closely to clinical phenotype observations, unlike phenotypes in OMIM or the HPO database which collect phenotypes across multiple cases. We randomly selected 500 variants each from a distinct gene along with their associated phenotypes from PAVS.

For each of the benchmark sets, we generated a set of pairs (*G, P*) of a list of genes *G* = (*G*_1_, …, *G*_*n*_) and a set of phenotypes *P* = (*P*_1_, …, *P*_*m*_). The phenotypes are identical to the phenotypes from the benchmark sets (which contain genotype– phenotype relations); the list of genes *G* contains the causative gene (i.e., the genotype mapped to the underlying gene) and a set of genes randomly chosen either from all human genes or from all genes with a genotype in the benchmark set. We vary the size of the gene set *G* by randomly choosing different numbers of genes to add; the cardinality of *G* ranges from 5 to 100 (cardinalities 5, 25, 50, 75, 100).

### 2.2 Baseline methods

We evaluated several state-of-the-art methods for phenotype-based gene prioritization, all implemented in the Exomiser [35] system. We use three main methods as baseline: ExomeWalker [36], PHIVE [37], and PhenIX [38], as well as their weighted combination (labeled “Exomiser score”). Exomiser uses phenotypes in the form of HPO terms as input and, because it is designed for ranking variants in whole exome or whole genome sequencing, it outputs a ranked list of variants. We generate a random variant in each gene as input and ignore all variant-related scores produced by Exomiser in our evaluation.

The different algorithms implemented in Exomiser differ primarily in the type of background knowledge they employ [35]. The most basic algorithm, PhenIX, relies only on human phenotypes to rank candidate genes [38], and therefore does not provide a phenotype-based score or rank for genes that are not known as human disease genes. The PHIVE algorithm, the other hand, compares the input phenotypes to mouse model phenotypes and relies on cross-species phenotype integration and human–mouse orthology for ranking candidate genes [37]. ExomeWalker [36] employs protein–protein interaction networks and the guilt-by-association principle to rank candidate genes. The final Exomiser score is based on a logistic regression model that assigns weights to the individual scores and generates a combined score.

We utilized the default settings to execute the tools on the generated synthetic datasets from PAVS and ClinVar, inputting the acquired HPO codes for each variant. We assessed the gene scores for each prediction method, excluding variant scores, since our focus is on gene prioritization.

#### 2.2.1 Large Language Models Used

We used three LLMs as part of this study, GPT-3.5-Turbo [22] and GPT-4 [39] and the Falcon180B model [40]. GPT-3.5-Turbo is an instruction-following LLM with 20 billion parameters, trained on data up to January 2022. GPT-4 is a multi-modal instruction-following model; the model is commercially available as a blackbox model and no technical details are publicly known. We used GPT-4 trained with data up to April 2023. Falcon 180B is an LLM that is publicly available. Falcon 180B was trained on 3.5 trillion tokens primarily consisting of data from the RefinedWeb dataset [41]. Falcon contains 180 billion parameters. It is available as a base model and as a pre-trained model for conversations. We used the Falcon 180B-Chat version in our experiments, with training data up to November 2022.

We accessed GPT-3.5-Turbo and GPT-4 through the API provided by OpenAI Inc. We also used the Falcon 180B-Chat model and ran it using eight A100 80GB GPUs as recommended in the release notes. We restricted the model to return only tokens with high confidence by setting the do sample variable to false.

### 2.3 Prompt engineering

As part of our interaction with the LLMs, we designed a structured prompts to engage with the LLMs through their API. We followed the GPT best practice guidelines [42] to design our prompts. We wrote clear instructions by following the suggested tactics (e.g., ask the model to adopt a persona, use delimiters to clearly indicate distinct parts of the input, specify the output format) and evaluated each prompt on our benchmarking datasets.

Table 1 shows the prompts with which we experimented (Table A1 illustrates example prompts). Prompts Q1, Q2, and Q3 are zero-shot [43], while Q4 constitutes a one-shot, chain-of-thought prompt [43, 44] instructing the LLM specifically on how to perform the gene ranking. In Q1, we instruct the LLM to rank the provided gene list. In Q2, additional patient-related information, including sex and mode of inheritance, is provided. In Q3, the LLM is prompted to rank genes based on their function, expression site, and relevant animal models if there is insufficient information about the gene itself available. Lastly, Q4, a one-shot, chain-of-thought prompt precisely instructs the LLM on the ranking process and the required output format.

**Table 1.** Prompts Crafted.

| ID | Type | Prompt |
| --- | --- | --- |
| Q1 | zero-shot | A patient presented with these clinical symptoms: [phenotypes]. Rank these genes according to their association with the symptoms of the patient: [genes]. |
| Q2 | zero-shot | A [male/female] patient who is suspected of having a [mode of inheritance/genetic] disease, presented with these clinical symptoms: [phenotypes]. Rank these genes according to their association with the symptoms of the patient:[genes] |
| Q3 | zero-shot | A [male/female] patient who is suspected of having a [mode of inheritance/genetic] disease, presented with these clinical symptoms: [phenotypes]. Rank these genes according to their association with the symptoms of the patient: [genes]. In the case of insufficient information, still try to rank these genes by using function, site of expression or information from animal models. |
| Q4 | one-shot, chain-of-thought | Role: You are an automated ranking system. You take a set of patient signs and symptoms (phenotypes) as input, as well as a set of genes in which a likely pathogenic variant has been identified using a bioinformatics system. You return a ranked list of genes according to the likelihood of the damaging variant in the gene causing the phenotypes of the patient. To do the ranking, first identify if there is any knowledge about mutations in the gene causing the same or similar phenotypes as observed in the patient. Use information about disease and phenotypes, animal models, gene functions, and anatomical site of expression. Automatically rank all genes on the last rank if no evidence exists, and rank all other genes based on the likelihood of causing the phenotypes. Your ranked list should include only the user provided genes and not any other gene. Example: A [male/female] patient who is suspected of having a [mode of inheritance/genetic] disease, presented with these clinical symptoms: [phenotypes]. Rank these genes according to their association with the symptoms of the patient: [genes]. Assistant: "Ranked List:" 1. Gene1 2. Gene2 3. Gene3 ...<br>A [male/female] patient who is suspected of having a [mode of inheritance/genetic] disease, presented with these clinical symptoms: [phenotypes]. Rank these genes according to their association with the symptoms of the patient:[genes] |

To identify the prompt to use, we assessed GPT-3.5-turbo’s performance on GPCards using a selection of nine randomly chosen genes and one causative gene retrieved from GPCards. Table 2 shows the performance results of the different queries, including several combinations and variations of the queries. Q2+Q3 denotes the utilization of Q3 when Q2 fails, i.e., when the LLM does not rank a given set of genes based on Q2. The symbol Q2 *−*sign indicates substituting “symptoms” with “signs and symptoms”, whereas Q2 pheno *−*represents replacing “symptoms” with “phenotypes” in Q2. Q2-full gene names represents using full gene names instead of their symbols in Q2.

**Table 2.**
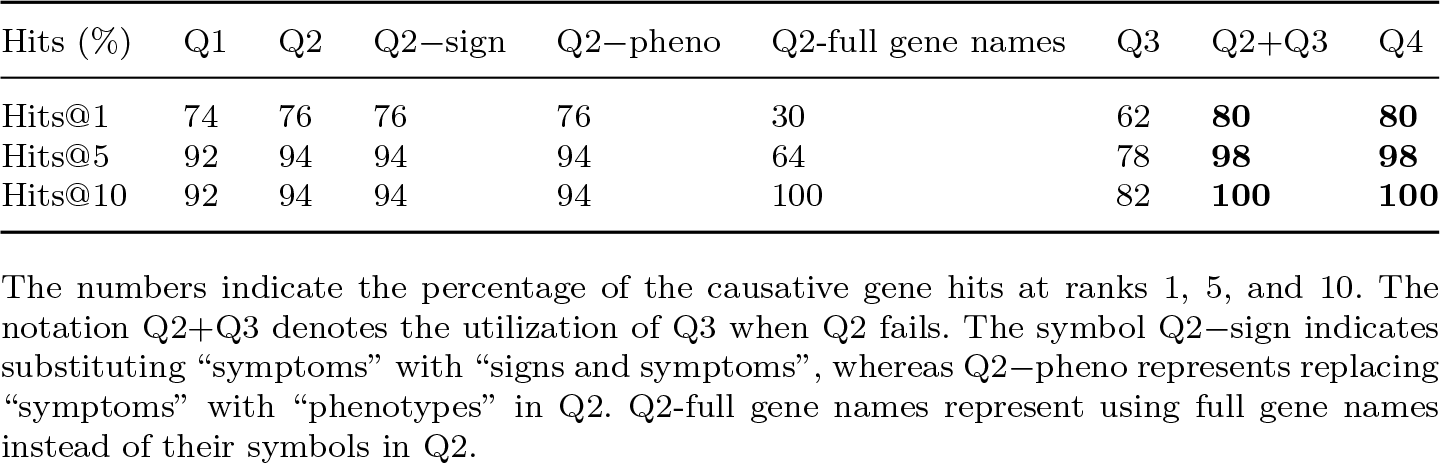
Evaluation of GPT-3.5-turbo with various prompts on GPCards.

| Hits (%) | Q1 | Q2 | Q2–sign | Q2–pheno | Q2–full gene names | Q3 | Q2+Q3 | Q4 |
| --- | --- | --- | --- | --- | --- | --- | --- | --- |
| Hits@1 | 74 | 76 | 76 | 76 | 30 | 62 | **80** | **80** |
| Hits@5 | 92 | 94 | 94 | 94 | 64 | 78 | **98** | **98** |
| Hits@10 | 92 | 94 | 94 | 94 | 100 | 82 | **100** | **100** |

The numbers indicate the percentage of the causative gene hits at ranks 1, 5, and 10. The notation Q2+Q3 denotes the utilization of Q3 when Q2 fails. The symbol Q2–sign indicates substituting “symptoms” with “signs and symptoms”, whereas Q2–pheno represents replacing “symptoms” with “phenotypes” in Q2. Q2–full gene names represent using full gene names instead of their symbols in Q2.

### 2.4 Rare disease cohort

We applied GPT-4 on 32 families presented at King Khalid University Hospital (KKUH) in Riyadh. Each family consisted of at least one individual with suspected genetic disease and multiple unaffected family members that provided blood samples at KKUH where DNA was extracted from blood. Using the extracted DNA, we constructed DNA libraries using a QIAGEN QIAseq FX DNA Library kit and sequenced each individual using an Illumina NovaSeq 6000 with an average coverage of 30x for each genome.

We used the bcbio-nextgen tool kit [45] and standard workflows to align the genomes to the GRCh38 human reference genome, to call variants using the GATK Haplotype caller [46], and genotype individuals.

After variant calling and genotyping, we filtered common variants (minor allele frequency less than 1%) using gnomAD (version 2.1.1) [47] and the 1,000 genomes project all population frequencies [48]. We then used the suspected mode of inheritance assigned by the clinical geneticist at KKUH based on the observed pattern of inheritance within the family and filtered variants by family pedigree using the slivar [10] software. We further removed variants not considered “impactful” by slivar tool, i.e., excluding synonymous and intronic variants [9].

### 2.5 Evaluation Metrics Used

We evaluated the performance of ranking genes using the area under the receiver operating characteristic curve (ROC AUC) [49], area under the precision–recall curve (AUPR), as well as recall (hits) at ranks 1, 5, and 10 indicating in how many cases the correct (causative) gene was retrieved at these ranks.

ROC AUC is calculated using

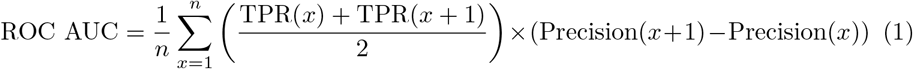

where 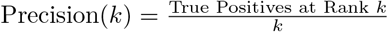 and *n* represents the total number of genes being ranked.

AUCPR quantifies the trade-off between precision and recall of a model across different thresholds:

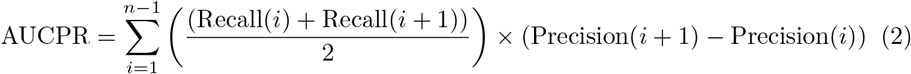

where Recall(*k*) and Precision(*k*) are precision and recall at rank *k*.

### 2.6 Availability of data

Primary or derived data from the families that were sequenced and analyzed is available only for researchers with access approved by the responsible IRB. Any requests for data access should be addressed to the Institutional Bioethics Committee at King Abdullah University of Science and Technology and the Institutional Review Board (IRB) at King Saud University.

### 2.7 Ethical approval declarations

1. Approval: This study was approved by the Institutional Bioethics Committee (IBEC) at King Abdullah University of Science and Technology under approval numbers 18IBEC10 and 22IBEC069, and the Institutional Review Board (IRB) at King Saud University under approval number 18/0093/IRB.
2. Compliance: All methods were carried out in accordance with the guidelines and regulations laid out by the institutional bioethics committees, the Declaration of Helsinki, and applicable laws and regulations governing research involving human subjects.
3. Informed consent: Informed consent was obtained from all participants or their legal guardians.

## 3 Results

### 3.1 LLMs accurately rank candidate genes

We applied LLMs to the problem of ranking genes based on a set of phenotypes associated with a Mendelian disorder. The input is a set of genes and a set of phenotypes, and the output is a ranked list of genes, with the gene most likely to be causative of the observed phenotypes ranked first. This evaluation reflects the use case where variants are already filtered by different evidence types or machine learning tools predicting pathogenicity, and the remaining variants have to be ranked based on whether the gene products they affect are likely involved in causing the observed set of phenotypes. In this ranking, only a few genes or gene products need to be considered.

We first applied LLMs on a dataset of genotype–phenotype relations derived from GPCards. GPCards contains variant-to-phenotype associations and describes the phenotypes associated with a genetic variant in natural language and does not rely on ontologies or structured vocabularies to characterize phenotypes. We also used GPCards to experiment with and optimize how to interact with the LLM through prompt engineering [50].

We designed a prompt in which we asked the LLM to rank a set of genes based on their likelihood of being involved in a set of phenotypes. The phenotypes we used in the prompt are taken from the phenotypes in the GPCards dataset, and one gene in the list of genes is the gene associated with the set of phenotypes in GPCards; the other genes are randomly chosen from all human genes. We additionally input the biological sex (if known) and the suspected mode of inheritance of the disease, if known. We evaluated the ranked list of genes generated by the LLMs as output, and determined where the positive gene (from the genotype-to-phenotype information in the GPCards database) is ranked.

Initially, we experimented with a set of “zero-shot” prompts [43] (see Materials and Methods) of the form:

~~~
A patient presented with these clinical symptoms: [phenotypes]. Rank these genes according to their association with the symptoms of the patient:[genes].
  and
A [male/female] patient who is suspected of having a [mode of inheritance/genetic] disease presented with these clinical symptoms: [phenotypes]. Rank these genes according to their association with the symptoms of the patient:[genes].
~~~

The information in square brackets is replaced by biological sex (if known, otherwise with an empty string), a comma-separated list of phenotypes, a comma-separated list of genes, and either the mode of inheritance (if known) or “genetic” if unknown.

The output of the LLM consists of a ranked list of genes, often with additional explanation in the form of one or two sentences that tries to explain why the gene is considered a relevant answer to the query (or why it is not). We ignored the explanations and evaluated the performance of the LLMs in ranking the “correct” gene.

We also experimented with other prompts, providing the LLM with a “reasoning” strategy about how it should identify relevant genes in the presence or absence of different types of information; we also provided an example of input and output to the model. This kind of interaction is a one-shot chain-of-thought prompt [44] because we provide an example and guide the model’s reasoning in ranking the genes based on phenotypes. The chain-of-thought (see Table 1) we designed is inspired by the different types of data that phenotype-based gene- and variant-prioritization methods like Exomiser [35] use.

To determine how well LLMs will rank genes based on their associations with a set of phenotypes when the number of genes between which it needs to discriminate increases, we increased the number of random genes we added to the causative one, from 4 to 99 (see Methods). We found that one-shot chain-of-thought prompting has the potential to improve over zero-shot prompting and GPT-4 outperformed all of the LLMs tested (see Table 3). We repeated each experiment five times to determine variance in ranking results and found that the ranks are highly consistent when ranking genes and results generally reproducible (Table B2).

**Table 3.**
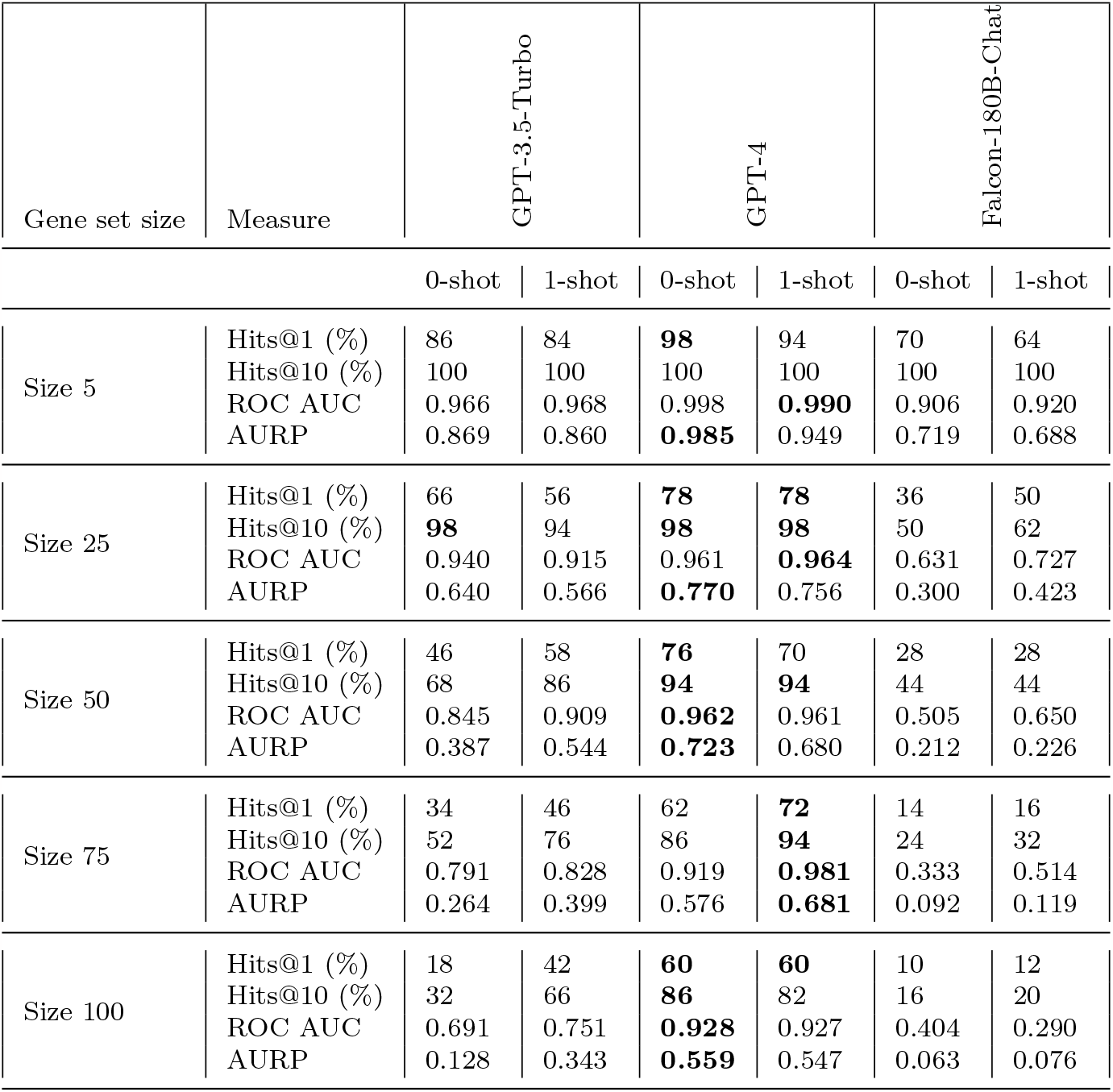
Evaluation on GPCards.

| Gene set size | Measure | GPT-3.5-Turbo |  | GPT-4 |  | Falcon-180B-Chat |  |
| --- | --- | --- | --- | --- | --- | --- | --- |
|  |  | 0-shot | 1-shot | 0-shot | 1-shot | 0-shot | 1-shot |
| Size 5 | Hits@1 (%) | 86 | 84 | **98** | 94 | 70 | 64 |
|  | Hits@10 (%) | 100 | 100 | 100 | 100 | 100 | 100 |
|  | ROC AUC | 0.966 | 0.968 | 0.998 | **0.990** | 0.906 | 0.920 |
|  | AURP | 0.869 | 0.860 | **0.985** | 0.949 | 0.719 | 0.688 |
| Size 25 | Hits@1 (%) | 66 | 56 | **78** | **78** | 36 | 50 |
|  | Hits@10 (%) | **98** | 94 | **98** | **98** | 50 | 62 |
|  | ROC AUC | 0.940 | 0.915 | 0.961 | **0.964** | 0.631 | 0.727 |
|  | AURP | 0.640 | 0.566 | **0.770** | 0.756 | 0.300 | 0.423 |
| Size 50 | Hits@1 (%) | 46 | 58 | **76** | 70 | 28 | 28 |
|  | Hits@10 (%) | 68 | 86 | **94** | **94** | 44 | 44 |
|  | ROC AUC | 0.845 | 0.909 | **0.962** | 0.961 | 0.505 | 0.650 |
|  | AURP | 0.387 | 0.544 | **0.723** | 0.680 | 0.212 | 0.226 |
| Size 75 | Hits@1 (%) | 34 | 46 | 62 | **72** | 14 | 16 |
|  | Hits@10 (%) | 52 | 76 | 86 | **94** | 24 | 32 |
|  | ROC AUC | 0.791 | 0.828 | 0.919 | **0.981** | 0.333 | 0.514 |
|  | AURP | 0.264 | 0.399 | 0.576 | **0.681** | 0.092 | 0.119 |
| Size 100 | Hits@1 (%) | 18 | 42 | **60** | **60** | 10 | 12 |
|  | Hits@10 (%) | 32 | 66 | **86** | 82 | 16 | 20 |
|  | ROC AUC | 0.691 | 0.751 | **0.928** | 0.927 | 0.404 | 0.290 |
|  | AURP | 0.128 | 0.343 | **0.559** | 0.547 | 0.063 | 0.076 |

### 3.2 LLMs improve on ontology-based ranking methods

While our results on the GPCards dataset show that LLMs can rank genes based on a set of phenotypes specified in natural language, the majority of phenotype-based gene- or variant-prioritization methods rely on input specified in a formal language based on phenotype ontologies [17, 35, 51–53]. The use of an ontology removes ambiguity in phenotype descriptions and enables access to background knowledge contained in phenotype ontologies [20]. To compare the use of LLMs with established ontology-based ranking methods, we followed the same setup in ranking a set of genes and identifying the causative genes given a set of phenotypes, and we compared LLMs with the ontology-based tool Exomiser. Exomiser implements multiple different algorithms for prioritizing candidate genes based on different sources of information; it uses human phenotypes in the PhenIX algorithm [38], mouse model phenotypes in PHIVE [37], human and other model organisms phenotypes in hiPHIVE [37], and protein–protein interaction networks in ExomeWalker [36].

We used two databases of genotype–phenotype associations for our evaluation and comparison. The first is ClinVar, which is used widely to benchmark variant- and gene-prioritization methods. ClinVar contains associations of variants with diseases (specified using their OMIM identifiers); the OMIM diseases can then be mapped to their phenotypes using the HPO database [16]. For evaluation, we used only variants that have been added after the knowledge cut-off date (2 July 2023 – 7 October 2023) for GPT-4. While ClinVar is a comprehensive dataset of genotype–phenotype relations, it does not associate variants with phenotypes observed clinically but rather with the disorder. Therefore, we also used the PAVS database, a database of phenotype-associated variants in Saudi Arabia, which contains clinically-reported phenotypes and the associated variants. For both sets of variants, we followed a similar procedure as for our previous evaluation: we input the gene affected by the variant together with 4, 24, 49, 74, or 99 randomly chosen genes and asked the LLMs to rank the list of genes given the phenotypes. As GPT-4 was the best-performing model, we only evaluated GPT-4 on this task.

Table 4 shows the results when ranking genes based on ClinVar variants and phenotypes. We found that GPT-4 with a one-shot chain-of-thought query performs better than a zero-shot query, and that GPT-4 ranked genes better than all baseline methods when only a few genes were included in the list, but its performance dropped compared to Exomiser when more genes than 25 needed to be ranked.

**Table 4.** Evaluation on ClinVar.

| Gene set size | Measure | GPT-4 |  | PHIVE | PhenIX | ExomeWalker | Exomiser |
| --- | --- | --- | --- | --- | --- | --- | --- |
|  |  | 0-shot | 1-shot |  |  |  |  |
| Size 5 | Hits@1 (%) | 93 | **97** | 54 | 86 | 28 | 89 |
|  | Hits@10 (%) | 100 | 100 | 100 | 100 | 100 | 100 |
|  | ROC AUC | **0.991** | **0.991** | 0.902 | 0.962 | 0.834 | 0.967 |
|  | AURP | 0.944 | **0.972** | 0.605 | 0.865 | 0.395 | 0.889 |
| Size 25 | Hits@1 (%) | 82 | **85** | 38 | 83 | 6 | 84 |
|  | Hits@10 (%) | 94 | **97** | 88 | 90 | 70 | 91 |
|  | ROC AUC | 0.964 | **0.982** | 0.845 | 0.930 | 0.720 | 0.938 |
|  | AURP | 0.802 | **0.856** | 0.337 | 0.784 | 0.089 | 0.799 |
| Size 50 | Hits@1 (%) | 81 | 78 | 35 | 81 | 3 | **84** |
|  | Hits@10 (%) | 96 | **97** | 59 | 88 | 35 | 91 |
|  | ROC AUC | 0.971 | **0.977** | 0.833 | 0.930 | 0.761 | 0.936 |
|  | AURP | **0.806** | 0.786 | 0.280 | 0.761 | 0.053 | 0.789 |
| Size 75 | Hits@1 (%) | 71 | 74 | 34 | 79 | 4 | **82** |
|  | Hits@10 (%) | 82 | **96** | 43 | 86 | 27 | 86 |
|  | ROC AUC | 0.925 | **0.980** | 0.828 | 0.927 | 0.753 | 0.932 |
|  | AURP | 0.663 | 0.742 | 0.262 | 0.742 | 0.047 | **0.771** |
| Size 100 | Hits@1 (%) | 68 | 70 | 34 | 79 | 1 | **82** |
|  | Hits@10 (%) | 81 | **89** | 39 | 85 | 11 | 85 |
|  | ROC AUC | 0.911 | 0.929 | 0.822 | 0.920 | 0.690 | **0.930** |
|  | AURP | 0.622 | 0.668 | 0.256 | 0.733 | 0.020 | **0.766** |

In the case of ClinVar, we used the phenotypes from the HPO database as input for each ranking problem; these phenotypes are identical to the phenotypes associated with the causative gene in the database used by Exomiser, and this may bias the results. Therefore, we also used genotypes with their clinical phenotypes recorded in the PAVS database for evaluation. Table 5 shows the results. In the PAVS dataset, phenotypes do not match exactly with phenotypes in Exomiser’s genotype-to-phenotype database and GPT-4 performed better than all other methods in ranking genes, demonstrating that it is more robust to noisy phenotype descriptions than methods based on semantic similarity and explicit genotype-to-phenotype databases.

**Table 5.** Evaluation on PAVS.

| Gene set size | Measure | GPT-4 |  | PHIVE | PhenIX | Exome Walker | Exomiser |
| --- | --- | --- | --- | --- | --- | --- | --- |
|  |  | 0-shot | 1-shot |  |  |  |  |
| Size 5 | Hits@1 (%) | 93 | **94.80** | 48.40 | 78.80 | 22.40 | 79.60 |
|  | Hits@10 (%) | 100 | 100 | 100 | 100 | 100 | 100 |
|  | ROC AUC | 0.989 | **0.991** | 0.863 | 0.930 | 0.788 | 0.927 |
|  | AURP | 0.943 | **0.956** | 0.538 | 0.788 | 0.335 | 0.771 |
| Size 25 | Hits@1 (%) | 75.40 | **77.80** | 31.40 | 69.60 | 4.60 | 65.60 |
|  | Hits@10 (%) | 97.80 | **99.00** | 75.60 | 82.60 | 57.40 | 82.60 |
|  | ROC AUC | 0.970 | **0.979** | 0.777 | 0.885 | 0.676 | 0.880 |
|  | AURP | 0.753 | **0.784** | 0.270 | 0.656 | 0.075 | 0.618 |
| Size 50 | Hits@1 (%) | 66.40 | **68.40** | 27.80 | 62.20 | 2.80 | 57.20 |
|  | Hits@10 (%) | 91.00 | **95.2** | 47.20 | 82.00 | 34.80 | 80.20 |
|  | ROC AUC | 0.949 | **0.973** | 0.769 | 0.881 | 0.713 | 0.875 |
|  | AURP | 0.639 | **0.677** | 0.219 | 0.586 | 0.046 | 0.540 |
| Size 75 | Hits@1 (%) | 59.80 | **61.80** | 26.00 | 55.60 | 3.00 | 51.60 |
|  | Hits@10 (%) | 83.40 | **91.40** | 37.80 | 82.00 | 21.80 | 79.40 |
|  | ROC AUC | **0.992** | 0.960 | 0.765 | 0.879 | 0.704 | 0.872 |
|  | AURP | 0.553 | **0.595** | 0.197 | 0.524 | 0.037 | 0.483 |
| Size 100 | Hits@1 (%) | 56.40 | **57.80** | 22.40 | 51.20 | 1 | 58.60 |
|  | Hits@10 (%) | 77.20 | **84.80** | 33.60 | 81.20 | 12.80 | 78.40 |
|  | ROC AUC | 0.895 | **0.937** | 0.756 | 0.874 | 0.658 | 0.870 |
|  | AURP | 0.500 | **0.539** | 0.178 | 0.481 | 0.019 | 0.445 |

### 3.3 LLMs reveal candidate genes for undiagnosed cases

We assessed the performance of LLM-based ranking of genes using a cohort of 32 families each with at least one individual with undiagnosed likely genetic disease. All affected individuals were seen at King Khalid University Hospital (KKUH) in Riyadh, Saudi Arabia, and whole genome sequencing was performed for affected individuals and their family members (see Methods). Neither genetic nor phenotype data for these families is publicly available or published; consequently, these cases represent a challenging “unseen” test case for the utility of LLMs in identifying causative genes in rare genetic diseases. The variants identified after whole genome sequencing were filtered by family pedigree, and suspected mode of inheritance, and allele frequency to retain only rare variants, and filter for potentially impactful variants (see Methods). After these filtering steps, the number of variants left in affected individuals ranged between one and 215 (mean 51.90), and these variants affected between one and 161 (mean 37.97) genes.

We used the genes with a potentially impactful variant after all filtering steps as the list of genes to rank, and the phenotypes observed clinically for each family as phenotypes, and used either the zero-shot or single-shot chain-of-thought prompt evaluated earlier. Based on our performance evaluation, we applied only GPT-4 to this cohort.

All 32 families underwent a detailed analysis to assess the biological and clinical plausibility of the top five candidate gene predictions. We assessed whether the top candidate from either the zero or few shot approaches had, in the opinion of two experts, a likelihood or possibility of being the causative gene. This assessment, while expert, is inevitably subjective. It took into account existing evidence that the gene had already been associated with a closely related phenotypic description presented in the case in a genotype-to-phenotype database; evidence for loss or gain of function of a gene giving rise to at least two of the phenotypes or phenotype domains seen in the family (for example delay in speech acquisition was regarded as an example of developmental delay and therefore closely related); concordance of gene function or process, as described either by Gene Ontology [54] or in the literature, with the phenotypic description; phenotypic concordance with loss or gain of function of an ortholog in an experimental model; functional or etiological relationship between the phenotype of the patient and a gene–phenotype (e.g., vermis hypoplasia was regarded as closely associated with seizures/epilepsy or other neurodevelopmental disorders as the two are closely linked). Given the large variation in the severity and spectrum of disease manifestations in many rare diseases [55], it was important to assess biological plausibility rather than scoring precise and complete phenotype matches.

We assessed the top five ranked genes for each of the 32 families. Candidates were delivered for all but one case where GPT-4 failed to rank. Of these, 15 families received at least one gene with a plausibility score of 4 or 5, and 23 genes were deemed plausible a total of 155 scored. Candidate genes had 212 OMIM Phenotype-Gene relationships (Phenotype-MIM number; some genes have more than one Phenotype-MIM record) but only 15 candidates had these previously asserted associations scored as the most plausible candidate. This was either due to very partial concordance of phenotypes – e.g. no more than one phenotype in common, irrelevant or discordant phenotypes, or differing modes of inheritance in combination with one of the above conditions. So a candidate with only one of the phenotypes of the OMIM allele recapitulated, and a recessive rather than dominant mode of inheritance would be regarded as a worse match. However, in the case of a good phenotype concordance and a discordant mode of inheritance, a lower match score was awarded on the premise that some alleles with different inheritance patterns might not have been previously described; generally scored a 4 or 3. There was no clear relationship between the quality of the explanation and the plausibility of the candidate (see below).

### 3.4 Explanations, hallucinations, and reproducibility

One of the advantages of LLMs in variant- and gene-prioritization is that they can not only perform the ranking of genes but can also provide explanations for the ranks assigned. However, due to the statistical nature of LLMs, they may also provide output that is factually incorrect, irrelevant, or inconsistent; we collectively refer to these outputs as “hallucinations”.

The first type of hallucination we observed was when the ranked list of genes included genes that were not specified as input, omitted genes that were provided as input, or contained duplicates (Table 6). We found that all LLMs we evaluated would often (in up to 56% of cases) remove genes from the list to rank, therefore not ranking all genes provided as input. Less frequently, LLMs also added new genes to the list to rank. Overall, we found that GPT-4 was more reliable than the other LLMs we evaluated, with the lowest number of hallucinations (both removing or adding genes from the list to rank), and that prompts where phenotypes use structured, ontology-based input instead of free-text were less prone to hallucinations than free-text input (Table 6).

**Table 6.** Hallucination results.

| Model and Prompt | GPCards |  | PAVS |  | ClinVar |  |
| --- | --- | --- | --- | --- | --- | --- |
|  | missing | extra | missing | extra | missing | extra |
| GPT-4 - zero shot | 42.00 | 00.80 | 14.48 | 01.84 | 15.40 | 01.60 |
| GPT-4 - one shot | 14.80 | 03.60 | 24.72 | 03.68 | 25.00 | 02.80 |
| GPT-3.5 turbo - zero shot | 56.80 | 00.80 | - | - | - | - |
| GPT-3.5 turbo - one shot | 46.40 | 30.40 | - | - | - | - |
| Falcon 180B - zero shot | 44.00 | 06.80 | - | - | - | - |
| Falcon 180B - one shot | 12.00 | 30.80 | - | - | - | - |

Missing indicates, the ranked list of genes missing one/more input genes. Extra indicates the ranked list of genes contains one/more genes that are not provided in the input list. The values are percentages obtained by using the results of all the prompts regardless of the gene size (5,25,50,75,100). We have 500, 250, and 2500 prompts for ClinVar, GPCards, and PAVS respectively.

We observed a second type of hallucination in the explanations that LLMs are generated for ranking certain genes. Hallucinations included those that gave an inappropriate response, ones that gave an irrelevant response, and the ones that seemed true and were convincing, but had no basis in fact. The latter kind of hallucination is potentially of the highest concern because it is challenging to detect and usually, requires expert review of available sources and literature.

We manually reviewed the quality of the explanation given by the LLM for its choices of the top five gene candidates for 32 families from our rare disease cohort. In some cases, the LLM gave a single global explanation which was often very general and factually correct but uninformative. In other cases, specific reasons were given for each gene.

We assessed the explanations provided in terms of their truthfulness, informativeness, completeness, and relevance. Truthfulness was assessed by whether the statement was factually true and could be substantiated with facts or reasonable inferences from facts. Informativeness was assessed on how much useful, relevant, or novel information was conveyed by the explanation. Completeness describes whether the explanation provides a rationale for all aspects of the patient phenotype. Relevance was assessed by the degree to which the explanation for gene association was biologically or clinically relevant to the phenotypes. A summary of results using a scoring ranging from 0 (very poor by the above criteria) to 5 (excellent, or, in the case of candidate genes, a very plausible suggestion) is available as Supplementary Data (KSUFamilies.xlsx). Our results show that most explanations were factually correct (Truthfulness: mean: 3.8, SD: 1.9), although usually uninformative (Informativeness: mean: 2.2, SD: 1.6) and incomplete (Completeness: mean 1.8, SD: 1.4), and often not relevant to the phenotypes observed (Relevance: mean 2.1, SD: 1.8). We also ranked plausibility for the gene being causative of the phenotypes; overall, across the top five genes, plausibility had a mean of 1.6 (SD: 1.7). However, if we only consider the highest-scoring gene among the five genes we evaluated, the mean plausibility is 3.7 (SD: 1.2), and for 15 families out of 32, we identified at least one candidate scoring with a plausibility of 4 or 5.

While most explanations were truthful, there are some exceptions where we have been unable to identify evidence for the truth of the statements made. For example, in a family where the affected individual has the phenotypes proteinuria, focal segmental glomerulonephritis, hypertension, absent patellae, hypoplastic nails, limited range of motion of the knees, dysmorphic facial features, and growth parameters below the third centile, and a suspected dominant mode of inheritance, GPR107 is the second-ranked candidate gene and provided with the following explanation: “GPR107: This gene encodes a protein that is a member of the G protein-coupled receptor superfamily. This protein has been shown to be a receptor for the sugar glucose, and is widely expressed in the central nervous system. While not directly linked to the symptoms, it could potentially be involved due to its role in glucose metabolism.” It is true that GPR107 is a *G* protein-coupled receptor, and it may be involved in glucose metabolism through its action on glucagon physiology via its binding to neuronostatin [56]. However, we have been unable to find any evidence that it binds glucose. Therefore, while the overall assertion is largely true, the LLM has hallucinated a part of its explanation: GPR107 does not bind glucose. Furthermore, while there is a wide range of phenotypes reported, there is no clear common linkage to glucose metabolism and we consider this to be another type of hallucination, i.e., generation of an irrelevant response.

We observed a similar hallucination in another family where the affected individual has astigmatism, Legg-Perthes [57], intellectual disability, and short stature. The explanation given for the top suggestion (SMPD3) is: “This gene is associated with Legg-Perthes disease, a condition that affects the hip joint in children and can lead to short stature.” While it is true that, in principle, a hip disorder can lead to short stature, there is no discoverable link between SMPD3 and Legg-Perthes disease, and the disorder is primarily associated with Col2A1 (OMIM:150600 [58]). It is, however, true that loss of function of the mouse ortholog gives rise to disproportionate dwarfism [59], which is in principle a match. However, there are no behavioural or ocular phenotypes in these mice. In this case, the LLM has made a partial connection with the gene and the phenotypes, but the assertion that it is known to be involved in Legg-Perthes disease is hallucinatory. In a third type of hallucination, GPT-4 invented a syndrome, “TOR3A syndrome” with associated phenotypes bearing some relation to those of the patient. We can find no reference in the literature to “TOR3A syndrome” but TOR1A is associated with torsion dystonia [60]; OMIM: 128100. GPT-4 seems to have associated the phenotype of one gene with another closely related in name and function. We have seen this problem several times. For example, MYH4 and MYH14, where MYH14 is associated with autosomal deafness (OMIM: 600652) which is part of the patient phenotype, but MYH4, the given candidate, has no association with deafness that we can discover.

## 4 Discussion

We studied the use of LLMs for the task of gene prioritization-based on phenotypes, a task that has traditionally been thought to rely on structured background knowledge. Our results demonstrate that LLMs can perform as well or better than custom-built tools for this task. Phenotype-based methods for ranking candidate genes consist of two main components: a knowledge-base of genotype-to-phenotype relations, and a similarity measure [17]. Often, they also contain structured background knowledge about how phenotypes are related, usually in the form on a phenotype ontology [20], and use a similarity measure based on the background knowledge making it a semantic similarity measure [19]. To perform better than phenotype-based gene prioritization methods, LLMs need to be able to replace these two main components. LLMs obtain their background knowledge from literature; the content of most genotype-to-phenotype databases will at least to large parts be reported in the literature (for example in the form of clinical case report [61]), and from our results, we can observe, similar to results in other clinical domains [24], that LLMs are as good or better in extracting the relevant information. LLMs also seem to be able to compute similarity between phenotypes as well or better than the custom-built similarity measures in Exomiser, demonstrated in particularly when using clinical phenotype descriptions as input to the ranking model (Table 5).

LLMs are very flexible in the input they take; in particular, LLMs can use arbitrary text as input, and we demonstrated this in one of our datasets which is not based on the HPO as vocabulary. However, in all experiments, we used only natural language labels as input to the LLM whereas the baseline methods implemented by Exomiser use HPO codes as input. In the past, the biomedical research community has made significant efforts in designing and developing phenotype ontologies [62], and in particular the HPO [16] has been developed to enable interoperability and applications such as candidate gene prioritization. Our results demonstrate that structured phenotypes are not required for the task of candidate gene prioritization and may actually be a limiting factor in their success, either due to incomplete or inaccurate information in ontologies that leads to incorrect entailments [63], or due to limitations in how much information can be expressed by a formal language like HPO in contrast to natural language such as used as input to LLMs. While our results apply only to one of the many applications of phenotype ontologies, in the future, their use and benefit should be re-evaluated in particular regarding the cost of developing and maintaining ontologies versus the availability of highly developed LLMs able to perform many different tasks.

Our experiments also show that LLMs go beyond gene prioritization systems in that they can provide explanations for their results. Furthermore, LLMs also have the potential to refine and update ranking results interactively. The best use of LLMs may therefore be not as a simply ranking system but rather as an interactive diagnostic assistant. However, future work still needs to address “hallucinations” as well as ways to quantify uncertainty; knowledge graphs and ontologies may provide ways to solve the problem of hallucination [64, 65] by providing structured knowledge.

One potential limitation of our study is that the LLM models we evaluate were trained on text (i.e., literature articles) that report variants in our testing set. This is also a concern for the methods implemented in the Exomiser tool which relies on phenotypes from genes that were previously reported. We tried to control for this by including in our evaluation only variants after the knowledge cut-off date of GPT-4. However, in the future, a prospective study using LLMs for the diagnosis of genetic disease, based on our findings regarding variance in results, prompt design, or hallucinations should be designed. Such a prospective study could also investigate the use of LLMs for non-coding variants or structural variants which we did not consider here, and evaluate ways to include structured background knowledge for knowledge-enhanced learning.

## Data Availability

All data produced in the present work are contained in the manuscript and methods to produce all data are available online at https://github.com/bio-ontology-research-group/LLM_GenePrioritization

## Supplementary information

## Acknowledgments

We acknowledge support from the KAUST Supercomputing Laboratory. P.N.S acknowledges the support of the Alan Turing Institute.

## Declarations

### Ethics approval

#### Approval

This study was approved by the Institutional Bioethics Committee (IBEC) at King Abdullah University of Science and Technology under approval numbers 18IBEC10 and 22IBEC069, and the Institutional Review Board (IRB) at King Saud University under approval number 18/0093/IRB.

#### Compliance

All methods were carried out in accordance with the guidelines and regulations laid out by the institutional bioethics committees, the Declaration of Helsinki, and applicable laws and regulations governing research involving human subjects.

### Consent to participate

Informed consent was obtained from all participants or their legal guardians.

### Consent for publication

Informed consent was obtained from all participants or their legal guardians.

### Availability of data, materials, and code

https://github.com/bio-ontology-research-group/LLM_GenePrioritization

### Authors’ contributions

Ş.K. designed the prompts, conducted the GPT experiment, evaluated the results, filtered the VCF files for all KSU samples, and subsequently ranked their genes using GPT. Ş.K. also contributed to the initial draft of the manuscript. M. Abdelhakim prepared the libraries for all KSU samples for sequencing and participated in the manual analysis of gene prioritization results for KSU families. A.A. executed experiments using other state-of-the-art tools, generating VCF files for all KSU samples, and contributed to the evaluation of other methods. S.T. conducted experiments using Falcon180B-Chat and contributed to the preparation of the evaluation script. P.N.S. participated in the manual evaluation of the KSU families and the initial drafting of the manuscript. M. Alghamdi provided the KSU samples, their clinical phenotypes, and pedigrees. R.H. conceived the study, contributed to the prompt design, and participated in the initial drafting of the manuscript. All authors have reviewed and approved the final version of the manuscript.

## Appendix A Example Prompts

## Appendix B Observed variance in ranking results

**Table A1.** Example prompts.

| ID | Type | Prompt Example |
| --- | --- | --- |
| Q1 | zero-shot | A patient presented with these clinical symptoms: "Recurrent urticaria", "Recurrent abdominal pain", "Fatigue", "Fever", "arthralgia", "Lymphadenopathy", "Elevated circulating C-reactive protein", "Glomerulonephritis", "Elevated erythrocyte sedimentation rate", "Anemia". Rank these genes according to their association with the symptoms of the patient: "KCNJ4", "VEPH1", "AGO3", "TRG-CCC2-1", "ERG", "DNASE1L3", "KLRF1", "IGHV1-58", "NDE1P1", "LINC02927" |
| Q2 | zero-shot | A female patient who is suspected of having a genetic disease, presented with these clinical symptoms: "Recurrent urticaria", "Recurrent abdominal pain", "Fatigue", "Fever", "arthralgia", "Lymphadenopathy", "Elevated circulating C-reactive protein", "Glomerulonephritis", "Elevated erythrocyte sedimentation rate", "Anemia". Rank these genes according to their association with the symptoms of the patient: "KCNJ4", "VEPH1", "AGO3", "TRG-CCC2-1", "ERG", "DNASE1L3", "KLRF1", "IGHV1-58", "NDE1P1", "LINC02927" |
| Q3 | zero-shot | A female patient who is suspected of having a genetic disease, presented with these clinical symptoms: "Recurrent urticaria", "Recurrent abdominal pain", "Fatigue", "Fever", "arthralgia", "Lymphadenopathy", "Elevated circulating C-reactive protein", "Glomerulonephritis", "Elevated erythrocyte sedimentation rate", "Anemia". Rank these genes according to their association with the symptoms of the patient: "KCNJ4", "VEPH1", "AGO3", "TRG-CCC2-1", "ERG", "DNASE1L3", "KLRF1", "IGHV1-58", "NDE1P1", "LINC02927". In the case of not enough information, still try to rank these genes by using function, site of expression or information from animal models. |
| Q4 | one-shot, chain-of-thought | Role: You are an automated ranking system. You take a set of patient signs and symptoms (phenotypes) as input, as well as a set of genes in which a likely pathogenic variant has been identified using a bioinformatics system. You return a ranked list of genes according to the likelihood of the damaging variant in the gene causing the phenotypes of the patient. To do the ranking, first identify if there is any knowledge about mutations in the gene causing the same or similar phenotypes as observed in the patient. Use information about disease and phenotypes, animal models, gene functions, and anatomical site of expression. Automatically rank all genes on the last rank if no evidence exists, and rank all other genes based on the likelihood of causing the phenotypes. Your ranked list should include only the user provided genes and not any other gene. Example: A female patient who is suspected of having a genetic disease, presented with these clinical symptoms: "Recurrent urticaria", "Recurrent abdominal pain", "Fatigue", "Fever", "arthralgia", "Lymphadenopathy", "Elevated circulating C-reactive protein", "Glomerulonephritis", "Elevated erythrocyte sedimentation rate", "Anemia". Rank these genes according to their association with the symptoms of the patient: "NKAPP1", "EXD2", "ENRICH2", "PDS5B", "CAMK2G", "DNASE1L3", "PCDH19", "ACADVL", "TRAF6P1", "CYP2T3P". Assistant: "Ranked List:" 1. DNASE1L3 2. TRAF6P1 3. ACADVL 4. CAMK2G 5. PCDH19 6. ERICH2 7. PDS5B 8. EXD2 9. NKAPP1 10. CYP2T3P<br>A female patient who is suspected of having a genetic disease, presented with these clinical symptoms: "Recurrent urticaria", "Recurrent abdominal pain", "Fatigue", "Fever", "arthralgia", "Lymphadenopathy", "Elevated circulating C-reactive protein", "Glomerulonephritis", "Elevated erythrocyte sedimentation rate", "Anemia". Rank these genes according to their association with the symptoms of the patient: "KCNJ4", "VEPH1", "AGO3", "TRG-CCC2-1", "ERG", "DNASE1L3", "KLRF1", "IGHV1-58", "NDE1P1", "LINC02927" |

**Table B2.** Mean and Standard deviation values in different runs.

| Gene set size | Run1 | Run2 | Run3 | Run4 | Run5 | Mean | Standard deviation |
| --- | --- | --- | --- | --- | --- | --- | --- |
| 5 | 0.990 | 0.986 | 0.992 | 0.990 | 0.992 | 0.990 | 0.0014 |
| 25 | 0.964 | 0.963 | 0.961 | 0.972 | 0.968 | 0.966 | 0.0028 |
| 50 | 0.961 | 0.966 | 0.965 | 0.961 | 0.979 | 0.966 | 0.0127 |
| 75 | 0.981 | 0.974 | 0.964 | 0.971 | 0.977 | 0.973 | 0.0028 |
| 100 | 0.927 | 0.948 | 0.947 | 0.950 | 0.909 | 0.936 | 0.0127 |

The runs are conducted using GPT-4 with the one-shot chain-of-thought prompt on GPCards, considering genes of different sizes. The results include ROC AUCs obtained from each run, as well as the corresponding mean and standard deviation values.

